# Health Disparities and COVID-19: A Retrospective Study Examining Individual and Community Factors Causing Disproportionate COVID-19 Outcomes in Cook County, Illinois, March 16-May 31, 2020

**DOI:** 10.1101/2020.08.21.20179317

**Authors:** Larissa H Unruh, Sadhana Dharmapuri, Xia Yinglin, Kenneth Soyemi

## Abstract

**Background:** Early data from the COVID-19 pandemic suggests that the disease has had a disproportionate impact on communities of color causing higher infection and mortality rates within those communities.

**Methods:** This study used demographic data from the 2018 US census estimates, mortality data from the Cook County Medical Examiner’s office, and testing results from the Illinois Department of Public Health to perform both bivariate and multivariate regression analyses to explore the role race plays in COVID-19 outcomes at the individual and community levels.

**Results:** Principal findings show that: 1) while Black Americans make up 22% of Cook County’s population, they account for 36% of the county’s COVID-19 related deaths; 2) the average age of death from COVID-19 is seven years younger for minorities compared to Non-Hispanic White (White) decedents; 3) minorities were more likely than Whites to have seven of the top 10 co-morbidities at death; 4) residents of predominantly minority areas were twice as likely to test positive for COVID-19 (p = 0.0001, IRR 1.94, 95% CI 1.50, 2.50) than residents of predominantly White areas; and 5) residents of predominantly minority areas were 1.43 times more likely to die of COVID-19 than those in predominantly White areas (p = 0.03).

**Conclusions:** There are notable differences in COVID-19 related outcomes between racial and ethnic groups at individual and community levels. We hope that this study will scientifically illustrate the health disparities experienced by communities of color and help to address the underlying systemic inequalities still prevalent within our country.

## Background

On March 11, 2020, the World Health Organization officially declared the COVID-19 respiratory illness caused by the SARS-CoV-2 virus a pandemic. As of June 30, the virus had infected over 10.25 million people worldwide and caused approximately 507,000 deaths. The United States has become the epicenter of this pandemic with an incidence of 2.6 million, or 25% of the estimated global infections, and approximately 126,000, or 25% of the reported global deaths^1^.

Recent data from the CDC suggests that the burden of COVID-19 morbidity and mortality is not uniformly distributed throughout the US population^2,3^. People of color, especially those within Black and Latinx communities, are disproportionately affected by the disease. These disparities have been widely discussed by media outlets, medical journals, health associations, and acknowledged by political leaders^2,4–6^. The resulting attention has refocused public awareness on the health inequalities experienced by communities of color in the US. Despite the wide media coverage, the amount of objective research analyzing the disparate outcomes between racial and ethnic groups remains lacking.

In “Racial Health Disparities and Covid-19 – Caution and Context,” recently published in *The New England Journal of Medicine*, Chowkwanyun and Reed discuss the need for careful data analysis before making blanket statements about racial differences in COVID-19 morbidity and mortality^7^. In keeping with Chowkwanyun and Reed’s recommendations, the current study describes the demographic distribution of mortality and explores whether there is a causal relationship between race, neighborhood factors, and COVID-19 in Cook County, Illinois.

## Methods

We performed retrospective analyses to explore COVID-19 outcomes between March 16 and May 31, 2020. We used three data sources to create two master spreadsheets to investigate individual and community (zip code level) aspects of COVID-19 morbidity and mortality for people of color. First, the Cook County Medical Examiner’s website provided COVID-19 mortality data^8^. Data elements included decedent demographics, primary and secondary causes of death, and date and location of death. Second, the Illinois Department of Public Health (IDPH) provided zip code-level data about the number of COVID-19 tests and positive cases^9^. Third, the 2018 US Census Bureau’s 5-year estimate for Cook County, Illinois, provided zip code level demographic data including total population, population density, average household size, income, racial/ethnic group composition, poverty, language, and education data^10^.

Secondary comorbidities were aggregated for supplementary analysis. Decedent’s zip codes were converted to Zip Code Tabulation Areas (ZCTAs), which are spatial representations of US Postal Service zip code service areas^11^. Mortality data was then aggregated by ZCTA and merged with ZCTA-level census data to create the community-level data set. Minority was defined as any race or ethnic group other than Non-Hispanic White (White). Minority predominant ZCTAs were defined as those in which the minority population was greater than the White population. For the preliminary analysis we calculated excess mortality by racial/ethnic groups as the number of moralities above that expected if mortality was distributed evenly between races. We calculated and compared the average age at death for all decedents and the average age of death for every racial and ethnic group. Using those averages, we compared trends between the top 20 ZCTAs in Chicago for testing and mortality data and the top ZCTAs for poverty and household size.

Descriptive statistics for continuous variables were presented as means ± standard deviations (SDs). Categorical variables were presented as frequencies and proportions. All statistical tests were two sided, and p-values < 0.05 were considered statistically significant. Data was tested for normality by the Shapiro-Wilk test. For baseline characteristics, ANOVA was used to calculate parametric p-values, the Krukas-Willis test was used for non-parametric pvalues, and the Fisher’s exact test was used for categorical covariates. Wilcoxon rank-sum tests were used as appropriate for continuous variables. Chi-square tests was used for categorical variable analysis to determine the differences between two groups.

To explore the relationship between COVID-19 mortality and contributing neighborhood factors, we used generalized linear modeling with the number of deaths as the dependent variable modeled against independent covariates (risk factors). Generalized linear modeling is a family of models (e.g., Poisson, negative binomial (NB), zero-inflated Poisson (ZIP), and zero-inflated negative binomial (ZINB)) that allow the linear regression model to be linked to the response variable by a link function using a backward elimination procedure. We then conducted a model comparison using criteria based on previous research^12–14^

In general, nested models are compared using a likelihood or a score test, while non-nested models are evaluated using the Vuong test. We compared the predictive performance of Poisson, NB, ZIP, and ZINB models using various indices, such as likelihood ratio, Akaike’s information criterion (AIC), Bayesian information criterion (BIC), standardized Pearson residual, and the standardized deviance residuals. In addition, differences between the predicted and actual counts were compared based on the mean squared error (MSE) performance measure. To compare the differences of the top 10 comorbidities among White, Black, Latinx, and other races, we fitted a generalized logit model using White as the reference. The figures were created using Microsoft Excel™ and statistical analysis was performed using SAS version 9.4 (SAS Inc., Cary, NC, USA).

## Results

Mortality case sample size included 3,762 COVID-19 related mortalities of which 201 cases without Cook County zip codes were removed. Of the 3,561 deaths, 1,281(36%) were Black, 1,387 (39%) were White, 656 (18%) were Latinx, and the rest were classified as Other. Males made up 2,060 (58%) of mortalities. Overall, mean (SD) age at death was 73.8 years(14.73). Hypertension was found to be the most frequent comorbidity and occurred in 2,403 (67%) of the decedents, 1,444 (40.5%) had diabetes (Table 1).

**Table 1:**
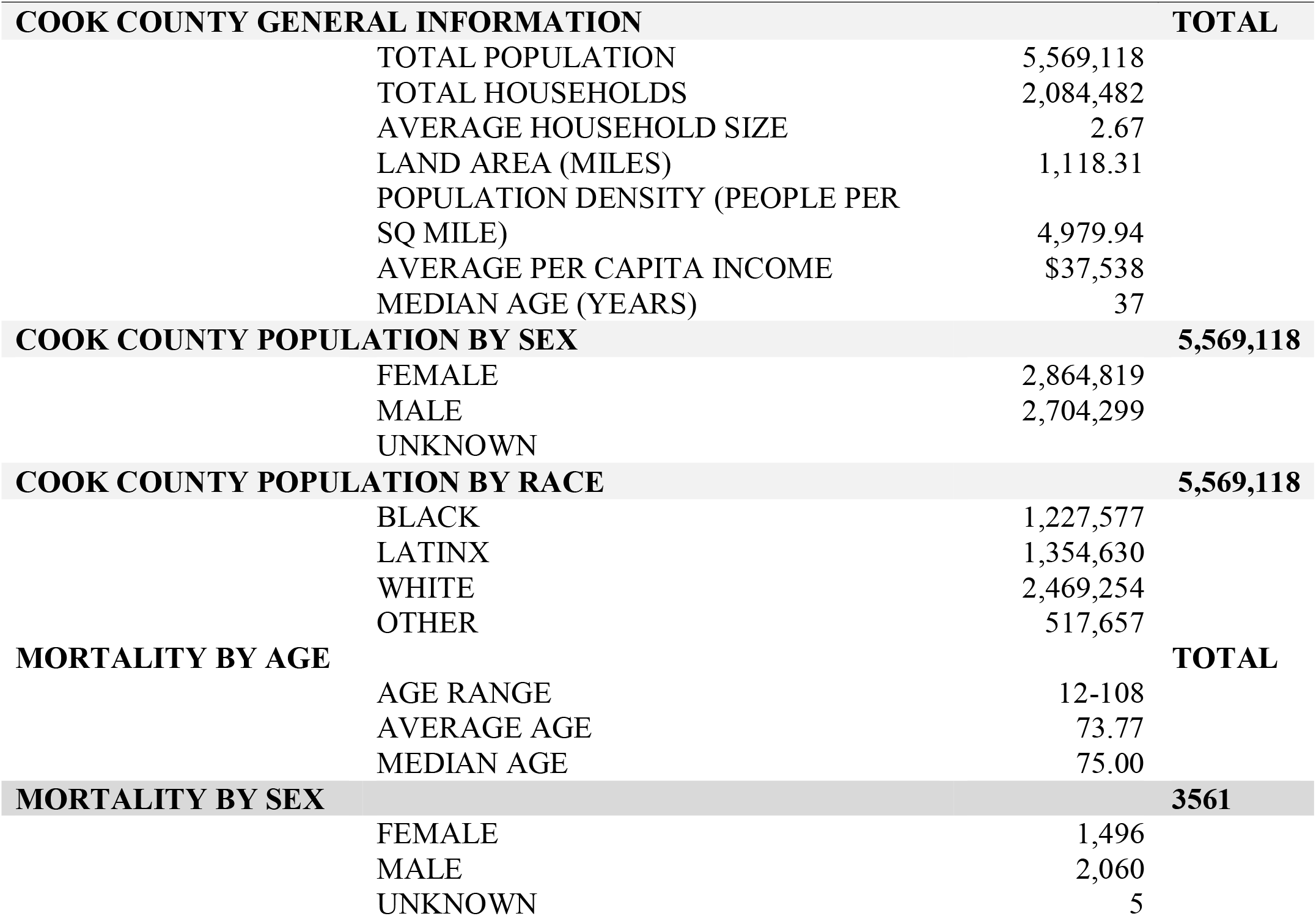
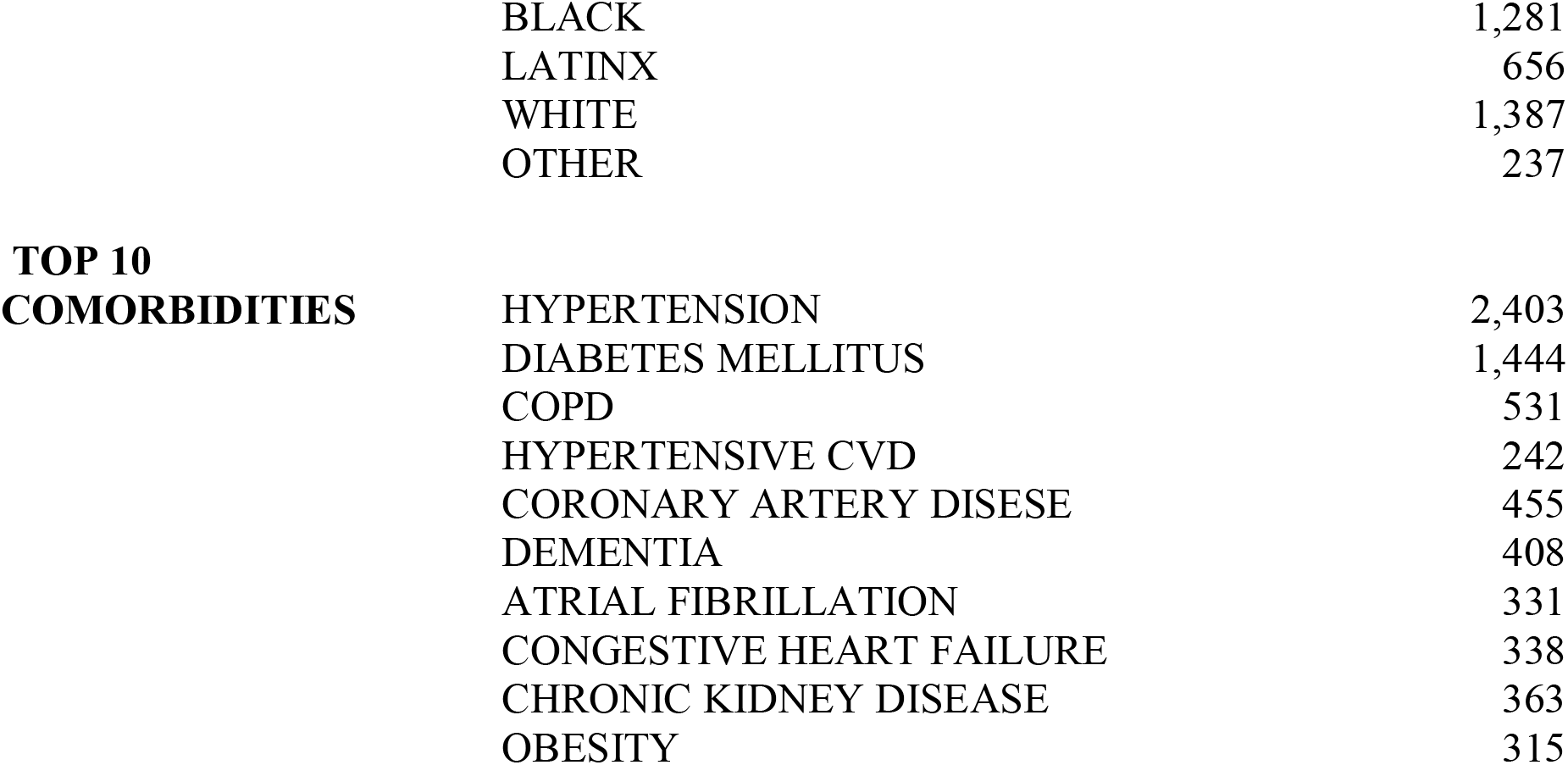
Population and COVID-19 Mortality: Cook County, IL: March 15-May 31, 2020.

| COOK COUNTY GENERAL INFORMATION |  |  | TOTAL |
| --- | --- | --- | --- |
|  | TOTAL POPULATION | 5,569,118 |  |
|  | TOTAL HOUSEHOLDS | 2,084,482 |  |
|  | AVERAGE HOUSEHOLD SIZE | 2.67 |  |
|  | LAND AREA (MILES) | 1,118.31 |  |
|  | POPULATION DENSITY (PEOPLE PER SQ MILE) | 4,979.94 |  |
| | AVERAGE PER CAPITA INCOME | \$37,538 | |
|  | MEDIAN AGE (YEARS) | 37 |  |
| COOK COUNTY POPULATION BY SEX |  |  | 5,569,118 |
|  | FEMALE | 2,864,819 |  |
|  | MALE | 2,704,299 |  |
|  | UNKNOWN |  |  |
| COOK COUNTY POPULATION BY RACE |  |  | 5,569,118 |
|  | BLACK | 1,227,577 |  |
|  | LATINX | 1,354,630 |  |
|  | WHITE | 2,469,254 |  |
|  | OTHER | 517,657 |  |
| MORTALITY BY AGE |  |  | TOTAL |
|  | AGE RANGE | 12-108 |  |
|  | AVERAGE AGE | 73.77 |  |
|  | MEDIAN AGE | 75.00 |  |
| MORTALITY BY SEX |  |  | 3561 |
|  | FEMALE | 1,496 |  |
|  | MALE | 2,060 |  |
|  | UNKNOWN | 5 |  |

|  |  |
| --- | --- |
| BLACK | 1,281 |
| LATINX | 656 |
| WHITE | 1,387 |
| OTHER | 237 |

**TOP 10 COMORBIDITIES**
|  |  |
| --- | --- |
| HYPERTENSION | 2,403 |
| DIABETES MELLITUS | 1,444 |
| COPD | 531 |
| HYPERTENSIVE CVD | 242 |
| CORONARY ARTERY DISESE | 455 |
| DEMENTIA | 408 |
| ATRIAL FIBRILLATION | 331 |
| CONGESTIVE HEART FAILURE | 338 |
| CHRONIC KIDNEY DISEASE | 363 |
| OBESITY | 315 |

Mean age at death was lower for minorities compared with white. Minority decedents had higher prevalences of diabetes, obesity, hypertension, coronary heart disease, and chronic kidney disease etc. compared with white decedents. Univariable analysis showed that the proportions of minority versus white were statistically significant for all the comorbidities minus hypertension and chronic kidney disease. Overall, the proportion of minorities with one or more co morbidity ranged from 41% to 67.7 % compared with 33% to 51.2% for whites. For both minorities and whites, males recorded higher proportion of deaths 1282 of 2173 (59%) vs 777 of 1387 (56%) respectively (Table 2).

**Table 2:**
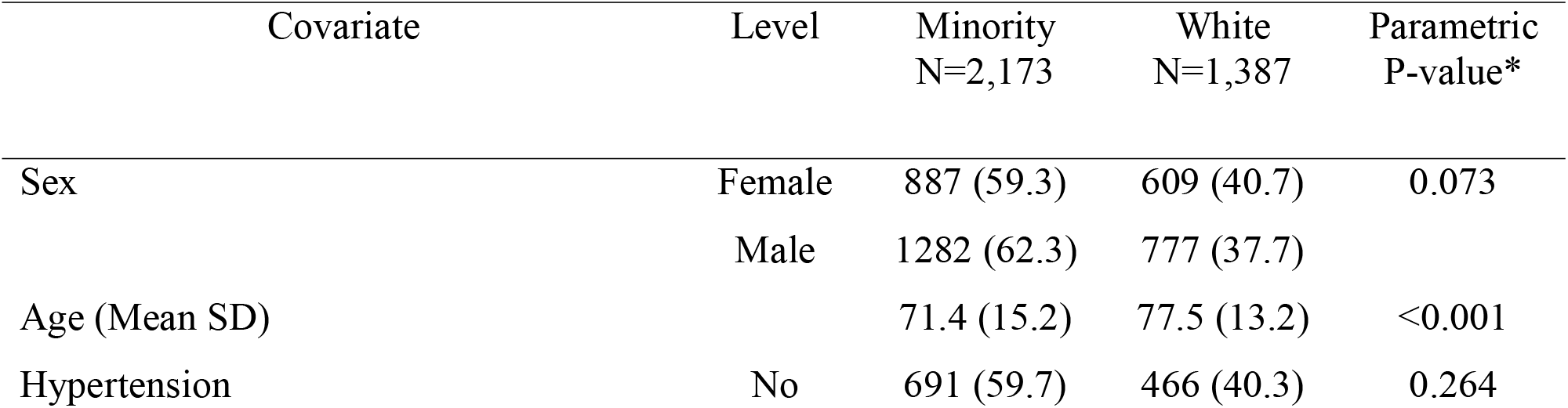
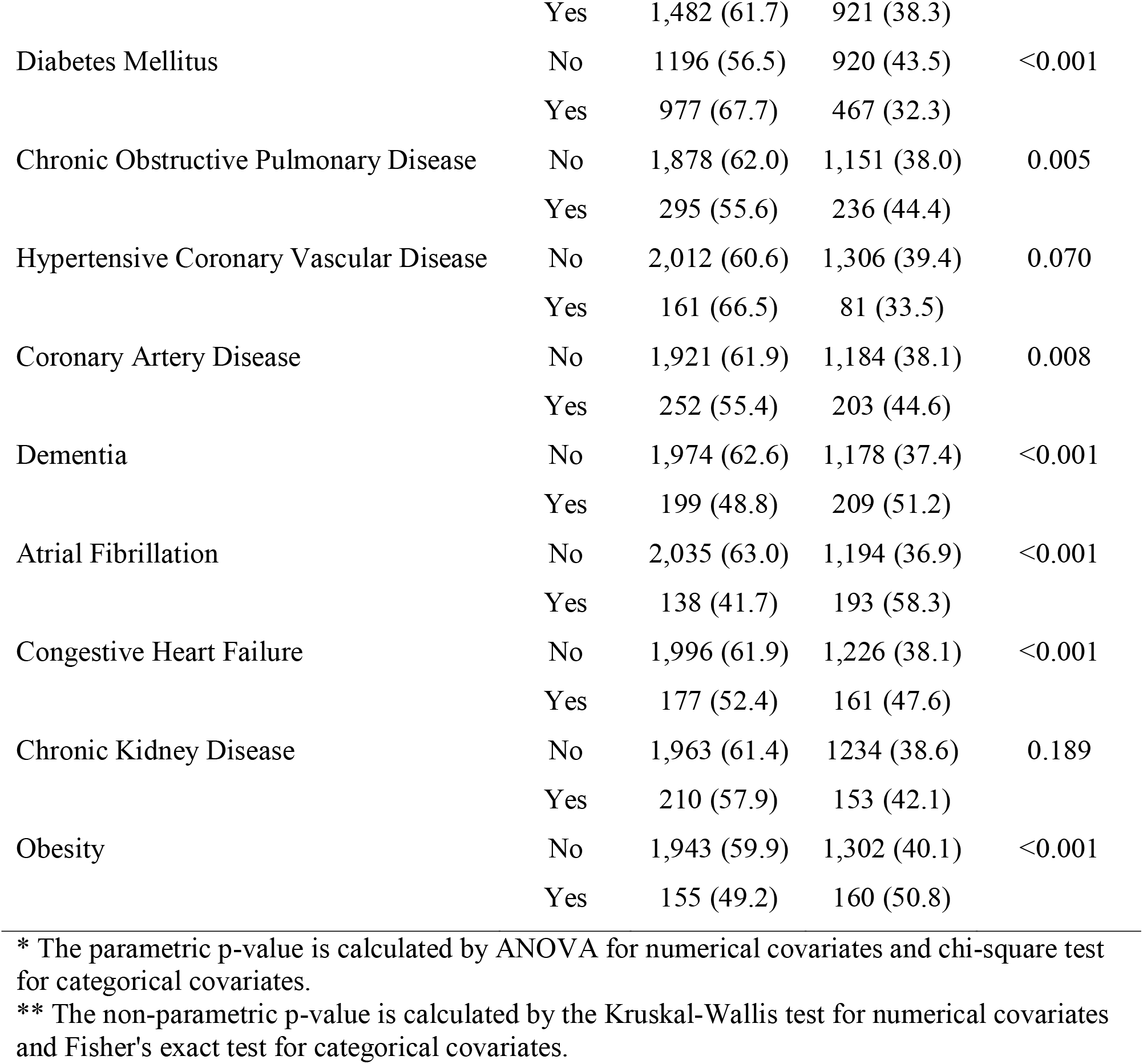
White vs. Minority Mortality: Cook County, IL, March 15 – May 31, 2020.

The ZCTA data set included 176 Cook County ZCTAs. Of the top 20 ZCTAs with the highest rates of COVID-19 infection, 15 (75%) were in predominantly minority areas. Residents of predominantly minority ZCTAs were two times as likely to test positive than people living in predominantly White ZCTAs [1.94 (1.50, 2.50) p = 0.0001]. We tested various models using NB regression which demonstrated that residents in predominantly minority ZCTAs were 58% or 1.4 times more likely to die compared with those in predominantly White areas. Of the risk factors analyzed, only the minority ZCTA variable was significantly associated with the number of COVID-19 deaths [1.42 (1.02, 1.97) p = 0.0361]. Poverty, however, was marginally significant (p = 0.0691). (See Table 3)

**Table 3:** Regression model outputs

| Model | IRR | 95% CI<br>Lower<br>Limit | 95% CI<br>Upper<br>Limit | Standard<br>Error | *P* value |
| --- | --- | --- | --- | --- | --- |
| Model 1 | 1.35 | 1.06 | 1.72 | 0.12 | 0.02* |
| Model 2 | 1.94 | 1.50 | 2.51 | 0.13 | <.00* |
| Model 3 | 1.42 | 1.02 | 1.98 | 0.17 | 0.04* |
| Model 4 | 1.29 | 0.88 | 1.90 | 0.20 | 0.19 |
|  | 1.24 | 0.79 | 1.95 | 0.23 | 0.36 |
| Model 5 | 1.44 | 0.97 | 2.13 | 0.20 | 0.07* |
| Model 6 | 1.66 | 1.00 | 2.74 | 0.26 | 0.05* |

|  | Dependent Variable | Independent Variable(s) |
| --- | --- | --- |
| Model 1: | Number of Residents Tested | Minority Zip |
| Model 2: | Number of Positive Results | Minority Zip |
| Model 3: | Number of Deaths | Minority Zip |
| Model 4: | Number of Deaths | Minority Zip and Poverty |
| Model 5: | Number of Deaths | Poverty |
| Model 6: | Number of Deaths | Average Household Size |
| IRR: | Incident Rate Ratio |  |
| * | Statistically significant, $P < 0.05$ | |
| Minority Zip: | Zip codes where minority population > white |  |
| Poverty: | Zip codes with > 20 percent of the population income is below the federal poverty level |  |

Using White decedents as a reference groups and simultaneously adjusting for other comorbidities, our multivariate analysis revealed several notable results. Of decedents with diabetes, Black individuals were 64% more likely to die of COVID-19 [Odds Ratio OR = 1.36(1.15, 1.62)], Latinx decedents were over two times as likely to die [OR = 2.10 (1.71. 2.59)], and other races were 63% more likely to die compared with White individuals. Of obese decedents, Black individuals were 49% more likely to die from COVID-19 [OR = 1.51 (1.08, 2.213)], Latinx individuals were again over two times as likely to die [OR = 2.17 (1.43, 3.28)], but other races were 80% less likely to die compared with obese White individuals [OR = 0.19 (0.02,1.43)]. Of decedents with hypertension, Black individuals were 62% more likely to die of COVID-19 [OR = 1.38 (1.15,1,65)], other combined races were 70% more likely to die [OR =1.29(0.86, 1.92)], but Latinx individuals were 38% less likely to die than White individuals [OR = 0.62 (0.5, 0.77)]. (See Table 4)

**Table 4:**
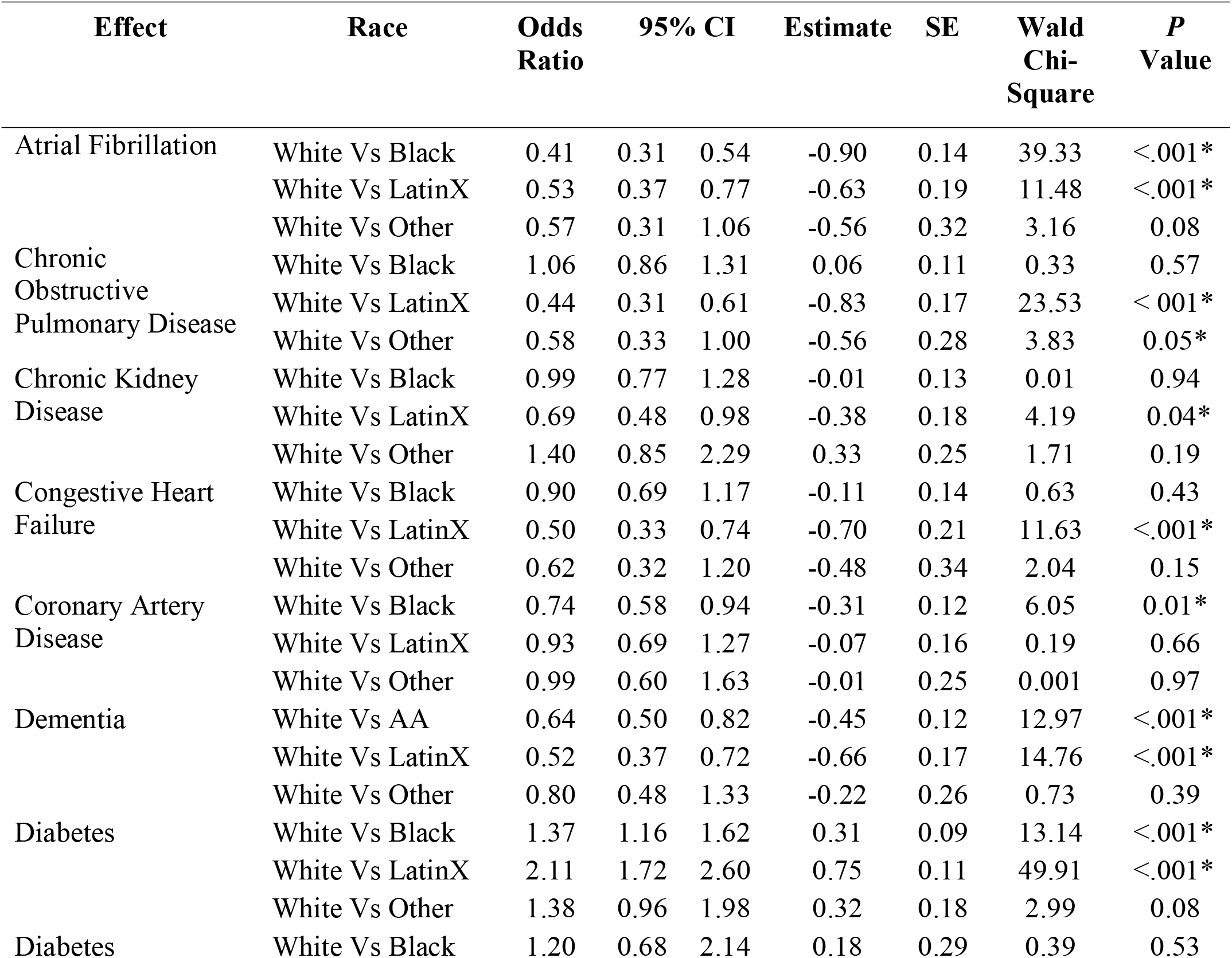
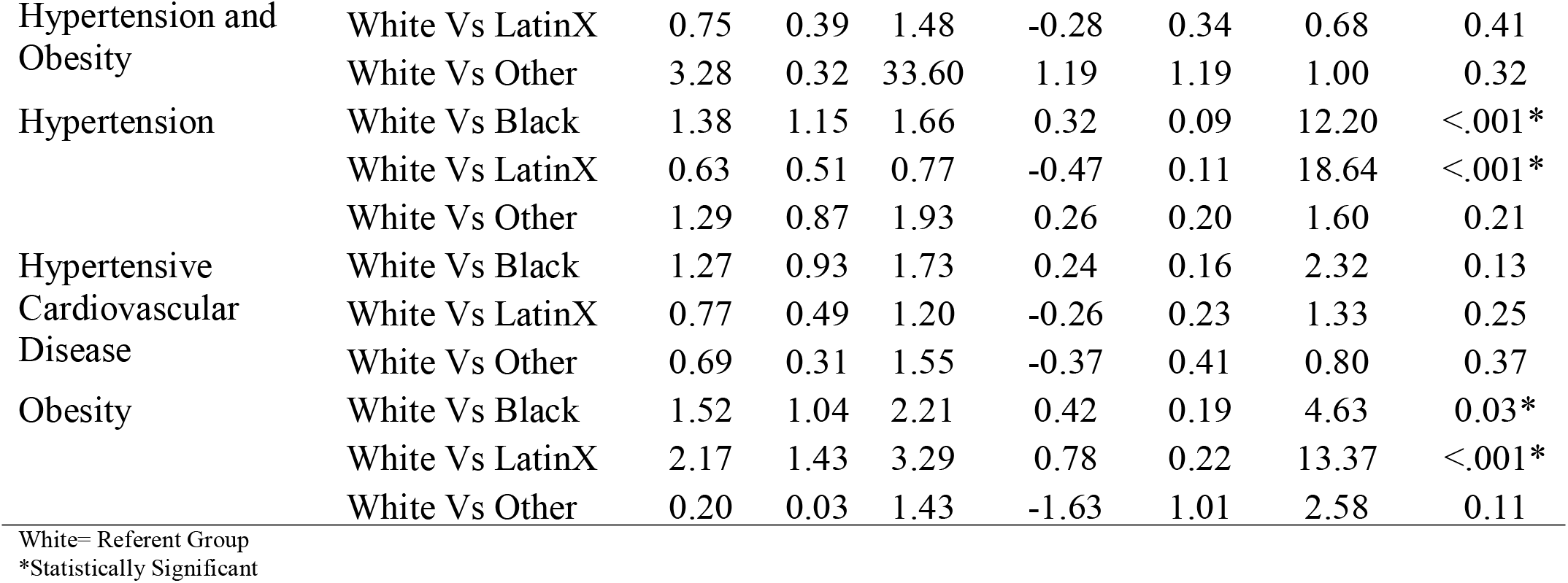
Co-Morbidities by Race

## Discussion

As of June 1, 2020, Cook County, Illinois, had identified over 78,000 positive SARS-CoV-2 cases and over 3,561 COVID-19 related deaths^15^. As with national COVID-19 trends, data from Cook County initially suggested that the burden of disease has fallen disproportionately upon communities of color. Although Black Americans make up 22% of Cook County’s population, they accounted for 36% of the county’s COVID-19 related deaths This finding suggests that the Black community is suffering a disproportionate amount of COVID-19 mortality. As outlined in other studies, our data showed that minorities are dying of COVID-19 at significantly younger ages than Whites (median minority age = 72, median White age = 79; p < 0.001). (Figure 1)

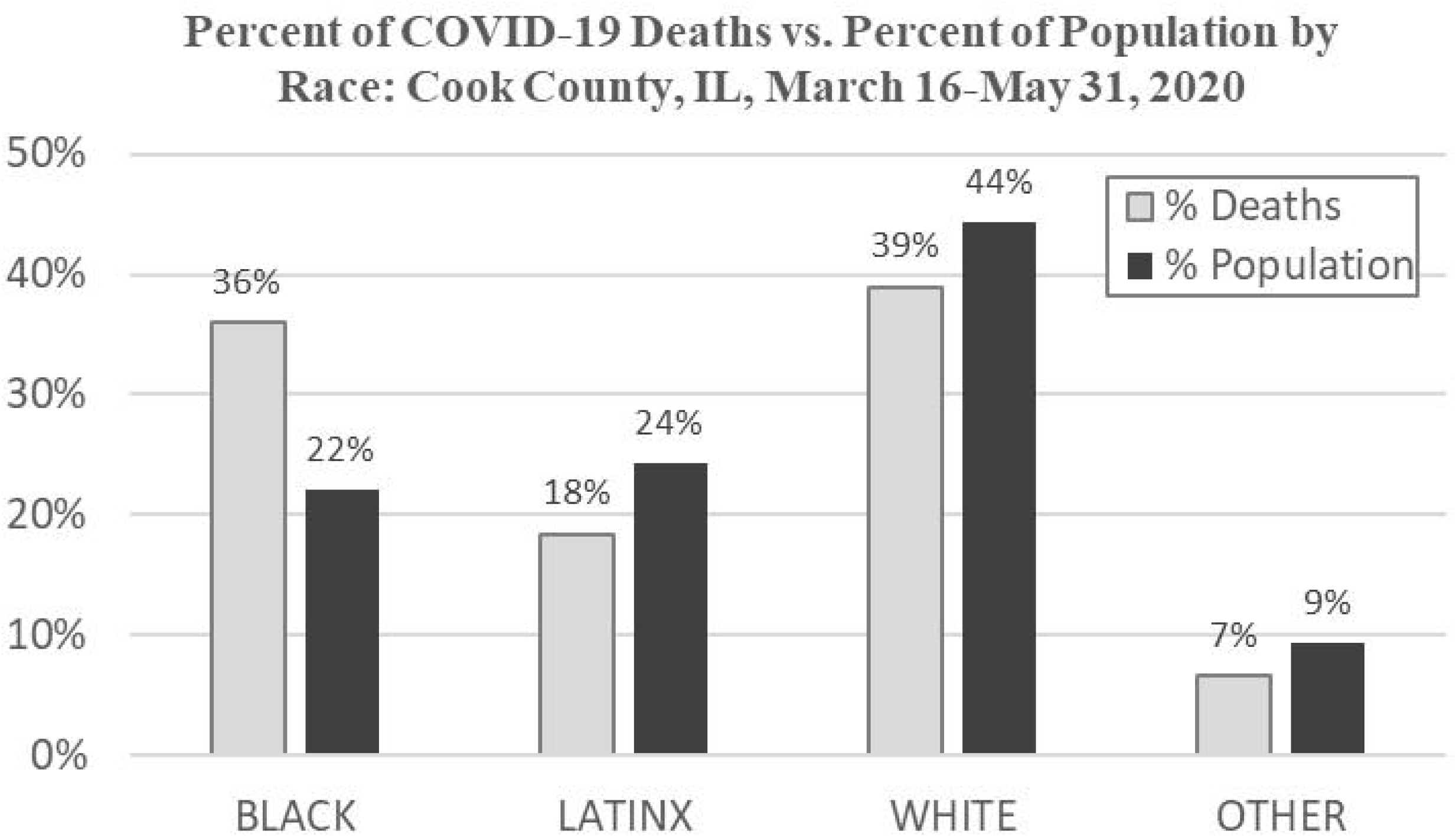
Figure 1:

We further explored the differences in average age at death by separating the data by race, which showed that Latinx decedents were, on average, more than 10 years younger than White decedents (Latinx = 67.3, White = 77.5)^16,17^. Additionally, minorities who died of COVID-19 were significantly more likely to have seven of the top 10 co-morbidities, including diabetes, chronic obstructive pulmonary disease, coronary artery disease, congestive heart failure, obesity, atrial fibrillation, and dementia.

Of the many reasons for COVID-19 disparities between races, we explored five: 1) lower socioeconomic status (SES), 2) increased comorbidities, 3) multigenerational living conditions, 4) high population density in predominantly Black and Latinx neighborhoods, and 5) minority overrepresentation in correctional facilities^6,18–21^.

### Socioeconomic Status

Khalatbari-Soltani *et al*. suggest that socioeconomic position plays a crucial role in COVID-19 outcomes and that these factors may be related to the differences in the COVID-19 burden observed between minority and White decedents^15^. While it may be true that certain factors, such as decreased access to nutritious food, less sanitary living conditions, or limited access to health care, affect all people with fewer economic resources, our analysis showed that poverty by itself was not a statistically significant factor for COVID-19 related death (p = 0.0691) indicating that there are likely other reasons for higher mortality among minorities. Of the factors explored, living in a predominantly minority ZCTA was the only statistically significant variable related to death from COVID-19. However, poverty may be confounded with living in a predominantly minority zip code. When poverty was analyzed in the same model as minority, neither variable was statistically significant (MZIP: p = 0.1880, POVERTY: p = 0.3592). In ZCTAs where over 20% of the population lived below the poverty line residents were 56% or 1.4 times more likely to die from COVID-19 than those in areas where less than 20% of the population lived below the poverty line (IRR = 1.44 (0.97, 2.13)].

### Increased Co-Morbidities

Other research has shown that people with certain co-morbidities (e.g., hypertension, diabetes, obesity) have suffered poorer COVID-19 health outcomes than people without underlying conditions^18^. As we compared comorbidities between ethnic/racial groups, we found that minority COVID-19 decedents had statistically significant chances of having seven of the top 10 comorbidities at the time of death. (Table 3).

### Multigenerational Living

In a study from Italy published in *Eurosurveillance*, Sjödin *et al*. explored factors necessary to mitigate the COVID-19 pandemic. The authors found that small household size was necessary for limiting the spread of the disease^22^. In Chicago, the areas with higher household density are primarily located in low density Black and Latinx neighborhoods. Our data showed that of the top 20 ZCTAs with the highest average number of people per household, 16 (80%) of them were in predominantly minority ZCTAs, and 12 (60%) of them were also in the top 20 for numbers of COVID-19 positive cases per 100,000 decedents. Our analysis confirms that larger household size is a statistically significant cause of COVID-19 mortality. Among all combined cases, for each one person increase in household size there is a 65.75% increase in mortality (p = 0.04).

### Densely Populated Urban Areas

Given that the virus spreads primarily through respiratory/droplet transmission, close contact with an infected individual is required for the virus to spread. It seems logical to assume that more highly populated ZCTAs have higher rates of infection and worse outcomes^23,24^. In Cook County, however, the more densely populated areas are the city center areas that are predominantly inhabited by wealthy, White individuals. Of the 20 ZCTAs with the highest population density, 12 are predominantly White, and 15 are above the average household income for Illinois. Additionally, these areas had lower infection rates with only one ZCTA in the top 20 for positive tests and lower mortality with two ZCTAs in the top 20 for mortality rate. At least in Chicago, infection and mortality rates seem to be more related to the density within households than overall population density.

### Overrepresentation in Correctional Facilities

Earlier in the pandemic, the Cook County Department of Corrections, one of the largest jail systems in the country, gained media attention about a large COVID-19 outbreak among both the facility’s employees and inmates^25–28^. As with many prisons, it has an over-representation of minority inmates. As of July 1, 2020, the Cook County Sheriff’s Department estimated that around 75% of Cook County inmates are Black^29^.

The demographics of the ZCTA that houses the Cook County Jail (60608) show that 55.4% of its inhabitants are male. This over-representation of the male population may be due to the presence of the prison system. The average number of people per household is 3.09, and the median age is 32.4 years. The population of the ZCTA is largely Latinx (50.7%) and Black(17.24%) with an average income of $21,447 (well below Illinois average household income). The COVID-19 data for the 60608 ZCTA shows that it ranks 26^th^ out of 176 for the most COVID-19 deaths and 9^th^ for the most positive cases in Cook County. Incidence analysis showed that of the deaths in the 60608 ZCTA, 24 of the 42 mortalities (57%) were Latinx, and 7 (16.6%) were Black. Therefore, although there is an over-representation of Black individuals within the jail, and the ZCTA had one of the highest incidents of COVID-19 positives, mortality data does not show that this ZCTA was an outlier area for COVID-19 mortality.

## Limitations

This study had several limitations. First, because we had data about individual mortality cases and broad ZCTA demographic data but lacked information about the demographics of those who did not die or who never tested positive for COVID-19, we were unable to perform a case-control analysis. Second, we used zip code demographic data as a proxy for an individual’s circumstances. Because that data was essentially an average, not everyone in that ZCTA was accurately represented. Third, because the study focused on Cook County, it may be generalizable only to large urban counties with high levels of racial and ethnic diversity. Fourth, census data is never fully able to capture the entirety of a population and may especially miss undocumented individuals. Fifth, because all death data was obtained from the Cook County Medical Examiner’s office, the completeness of that data depended on the Examiner’s reporting. Sixth, since the analysis relied on zip code level data, and zip codes do not remain constant over time, it was subject to the inherent issues associated with small area analyses. Seventh, we do not currently have data to indicate whether some ZCTAs with high mortalities had an overrepresentation of congregate care facilities such as long-term care or rehabilitation facilities which might skew the data.

## Conclusion

While we agree that caution is important when making statements about race and disease outcomes, our data indicate that the public health crisis created by inbuilt barriers within minority communities has contributed to the disproportionate mortality burden in Black and Latinx communities. It is time to not only to address the disparities that exist between racial/ethnic groups as they relate to COVID-19 but also the underlying structural issues that permit these disparities.

Our study shows that race, more than SES, is the only statistically significant factor for COVID-19 related mortality in Cook County. Other factors being equal, Black individuals are significantly more likely to die from COVID-19 overall, and Latinx individuals are dying at significantly younger ages than Whites. The question of why people of color suffer disproportionate COVID-19 mortality is more complicated than simply identifying that disproportionality. These data indicate that many of the deaths were not inevitable but were the byproducts of ingrained structural inequality resulting in diminished opportunities. We hope that the lessons from this study can help illuminate the persistent inequalities in our country so that, as a society, we can better address these issues. As we brace for a second wave of coronavirus outbreaks and a possible third wave, we must acknowledge who within our society suffers the most and find ways to focus our efforts to keep those populations safe and healthy.

## Data Availability

Data will be provided as needed

## Acknowledgments

Special thanks to Mary and Gail Unruh, Jeff Nogaj, and Dr. Mark Mycyk for their unwavering help and support.

